# Robust methylome analysis and tumour–normal classification in TCGA–COAD: a reproducible workflow

**DOI:** 10.1101/2025.10.16.25338162

**Authors:** Matthew Iwada Ekum, Christian Heumann, Eno Emmanuella Akarawak, Khalid Olajide Adekoya, Segun Isaac Oke, Adeyinka Solomon Ogunsnaya, Chongsheng Zhang, Olalekan Joseph Akintande, Joseph Adekunle Akinyemi, Isaac Ajiboye, Oluwafemi Daniel Amusa

## Abstract

Colorectal adenocarcinoma is caused in part by widespread epigenetic deregulation, yet the analysis of genome-wide DNA methylation of colorectal adenocarcinoma is complicated due to spatial correlation among CpG, multiscale patterns of differential methylation, and confounding cellular heterogeneity in bulk tissue. This study develops a simple yet effective framework that combines rigorous statistical modelling with modern machine learning-based prediction, on the Illumina 450K data from The Cancer Genome Atlas (TCGA) colorectal adenocarcinoma cohort. In our framework, differentially methylated regions (DMRs) were first detected using functional smoothing and permutation-based bump hunting, revealing both focal CpG island hypermethylation and broad hypomethylated domains spanning hundreds of kilobases. Next, we performed reference-based cell-type deconvolution and surrogate variable analysis (SVA) controlled immune/stromal admixture and hidden confounding effects, yielding well-calibrated single-site and region-level inference. Then, for tumour-status prediction, we empirically study the performance of classical Logistic Regression model against Random Forest, Gradient Boosting (XGBoost), and Feed-forward neural network; the results show that Logistic Regression achieves the lowest root-mean-square error and Brier score, reflecting its superior probability calibration. In general, our integrated framework provides biologically interpretable and highly predictive methylation signatures of colorectal cancer and offers a transferable baseline for future large-scale cancer epigenomics studies. Our code is publicly available at https://github.com/matekum/tcga-coad-methylation-ekum-2025a.

**Author summary:** We present a reproducible framework for analyzing genome-wide DNA methylation in colorectal cancer using public TCGA data. Our aim is twofold. We started by making single-site and regional findings statistically reliable by accounting for spatial correlation, hidden confounding, and cell-type mixtures; and second, we evaluate practical classifiers for tumour vs. normal prediction. We smooth effect sizes across neighbouring CpG sites and use permutation tests to detect differentially methylated regions, while surrogate variable analysis and reference-based cell-fraction estimates reduce unwanted variation. We then compare logistic regression with random forests, gradient boosting, and a neural network using principal component features. Across models, we observe near-perfect ranking of samples and strong probability calibration, with logistic regression performing best on calibration. All code, parameters, and figure scripts are openly available so others can reproduce and adapt the framework. Beyond colorectal cancer, the approach provides a template for robust methylome analysis and predictive modelling in other large-scale epigenomic studies.

## Introduction

Colorectal adenocarcinoma (COAD) is a leading cause of cancer-related morbidity and mortality worldwide. Aberrant DNA methylation is a hallmark of its pathogenesis, driving gene silencing, chromatin remodelling, and genomic instability. Genome-wide studies have revealed that epigenetic alterations occur across multiple genomic scales: focal hypermethylation at CpG islands, broad hypomethylated blocks spanning hundreds of kilobases, and large domains associated with higher-order chromatin organization [1, 2].

These multi-scale methylation patterns present substantial statistical challenges. First, CpG sites exhibit strong spatial correlation, meaning that neighboring sites tend to have similar methylation levels rather than varying independently. Second, methylation measurements are bounded between 0 and 1, which violates assumptions of standard linear models. Third, bulk tumour samples comprise heterogeneous mixtures of epithelial, stromal, and immune cells; consequently, differences in cell-type composition can confound true disease associations, leading to spurious findings.

Robust statistical inference therefore requires methods that simultaneously model multi-resolution methylation changes, control false discoveries under spatial dependence, and adjust for cellular heterogeneity [3, 4].

### From single-CpG testing to region-level inference

A *CpG* is a cytosine–phosphate–guanine dinucleotide (Fig. 1A). Dense clusters of these CpGs, termed CpG islands, frequently occur near gene promoters (Fig. 1B). When methylated, through the biochemical conversion of cytosine to 5-methylcytosine, these sites can repress gene transcription by reducing chromatin accessibility (Fig. 1C) [5, 6].

**Fig 1.**
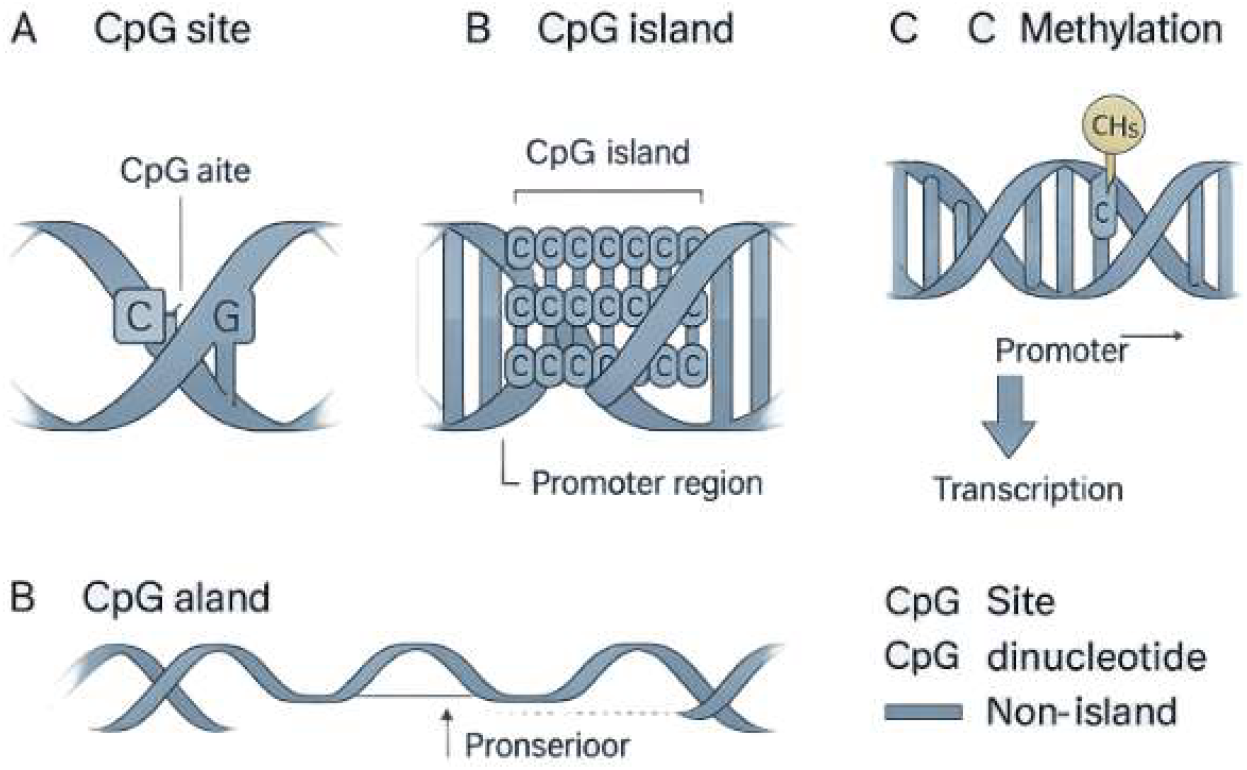
Schematic of CpG biology: (A) a CpG dinucleotide, (B) a CpG island near a promoter, and (C) 5-methylcytosine (CH_3_) at CpG sites and the potential repression of transcription. *Source: designed and generated by the authors*.

Classical statistical approaches test each CpG site individually using methods such as t-test or empirical Bayes linear models. These per-site tests provide a natural starting point and yield stable effect estimates in large methylation datasets. However, this site-by-site strategy has a fundamental limitation: tumour-associated methylation changes typically do not occur at isolated sites. Instead, they often extend across multiple neighboring CpGs in coordinated patterns. Testing sites individually fail to take advantage of this spatial structure and can miss broader regional changes.

To address this limitation, a two-stage approach that combines per-CpG modelling with region-level inference was developed [7, 8]. First, statistical models are fit at individual CpG sites to estimate methylation differences. Second, these effect estimates are smoothed along the genome and permutation-based “bump hunting” is applied to identify differentially methylated regions while controlling family-wise error rates. This strategy successfully detects both fine-scale changes at CpG-island shores (transition zones between CpG islands and bulk chromatin) and the broad hypomethylated blocks spanning hundreds of kilobases that characterise the colorectal tumour epigenome [9, 10].

### Controlling cellular heterogeneity

Single-cell epigenomic studies show that morphologically similar cells can differ markedly in chromatin state and gene expression [11–16]. Bulk tumour and adjacent normal tissues therefore contain mixtures of cell types with distinct methylomes. If unmodelled, such variation can mimic disease or age effects and inflate false discoveries. Reference-based deconvolution of leukocyte and epithelial fractions [17, 18] and surrogate variable analysis (SVA) [19] are now standard for removing these confounders. In the present study we incorporate estimated cell fractions directly in the design matrix and apply SVA to capture additional unmeasured variation, thereby improving both validity and reproducibility.

### Statistical modelling and machine-learning prediction

Genome-wide DNA methylation profiles are increasingly recognized as powerful biomarkers for cancer detection and patient stratification [20–25]. While high-performing classifiers such as gradient boosting, random forests, and deep neural networks can achieve excellent tumour–normal discrimination, clinical translation requires not only accuracy, but also well-calibrated risk estimates that accurately reflect true probabilities, as well as transparent interpretations that explain which genomic features determine such predictions [26–31].

To address these complementary objectives, we propose an integrated analytical framework comprising three components. First, we perform empirical-Bayes differential methylation analysis to identify both single-site changes and region-level alterations, providing mechanistic insight into the epigenetic alterations that characterize colorectal tumours. Second, we apply unsupervised dimension-reduction techniques, which are principal component analysis and hierarchical clustering, to visualize global methylation patterns and reveal sample structure independent of classification labels. Third, we conduct a rigorous predictive modelling benchmark that compares classical logistic regression with representative machine-learning approaches under nested cross-validation, ensuring unbiased performance estimates.

We used logistic regression with the standard logit link, providing calibrated probabilities and interpretable odds ratios that quantify the association between methylation features and tumour status. Moreover, we evaluate this interpretable baseline alongside more complex tree-based and neural network approaches to assess the trade-off between interpretability and predictive performance. This multi-layer strategy bridges mechanistic epigenetic discovery with clinically actionable prediction, integrating perspectives that previous studies have typically addressed in isolation.

### Related empirical evidence

Previous studies illustrate the need for such integration. [32] used TCGA 450K data to characterise age-related methylation changes in colorectal cancer but did not combine strict cell-composition correction, permutation-based DMR inference and machine-learning prediction. [33] performed multi-omics integration for biomarker discovery, and [34] combined methylation and fragmentomic profiles for cfDNA-based diagnosis, both reporting strong predictive accuracy but without region-level methylation modelling. [35] applied finite-mixture survival models to TCGA methylation profiles, focusing on prognosis rather than genome-wide DMR discovery or classification. Other related efforts span chromatin-regulator prognostic models [36], transcriptomic biomarkers [37], and integrative multi-omics recurrence signatures in endometrial cancer [38], as well as cfDNA methylation panels validated by qMSP [39]. These works highlight the potential of methylation and multi-omics biomarkers but rarely combine region-level inference with strict confounder adjustment and systematic comparison of multiple learners under nested cross-validation.

In this context, we present a unified, fully scripted R workflow for TCGA colorectal adenocarcinoma that delivers multi-resolution methylation inference and a carefully benchmarked machine-learning prediction pipeline. The framework provides bioinformaticians and biostatisticians with a reproducible large-scale methylation analysis toolkit, while giving cancer biologists and clinicians region-level maps and predictive models suitable for future diagnostic and translational studies.

## Materials and methods

### Data source and cohort

We conducted a secondary analysis of colorectal adenocarcinoma (COAD) from The Cancer Genome Atlas (TCGA). Illumina Infinium HumanMethylation450 (450K) array data and metadata were obtained via the Genomic Data Commons using `TCGAbiolinks`. After quality control (QC), the analytic cohort comprised 312 primary tumours and 38 solid-tissue normals. Raw IDATs were processed with background correction, NOOB normalisation, and probe filtering (cross–reactive probes, probes overlapping common SNPs, and sex-chromosome probes removed). Methylation levels were summarised as *β* and transformed to *M* = log_2_ {*β/*(1 −*β*)} for modelling [40, 41]. Implementation details, versioned package lists, and scripts are provided in the Supporting Information (S1 Methods).

### Preprocessing and exploratory analysis

Arrays were background-corrected and NOOB-normalised; poor-quality or non-finite values were handled by per-row medians where necessary. The exploratory plots included sample-wise density curves of *β* values, principal component analysis (PCA) on top-variance CpGs, and a heatmap of the top 1,000 most variable CpGs (row *z* scores). A representative heatmap is shown in Fig **??**.

### Statistical framework

#### Row-wise *z*-scores for the top 1,000 most variable CpGs

Let *X* ∈ [0, 1]^*p×n*^ denote the matrix of methylation *β*-values with CpG sites indexed by *i* = 1, …, *p* (rows) and samples by *j* = 1, …, *n* (columns). For each CpG *i*, let 𝒥_*i*_ = {*j*: *x*_*ij*_ is finite} and *n*_*i*_ = |𝒥_*i*_|.

#### Selection of CpGs by variability

Define the (empirical) row variance

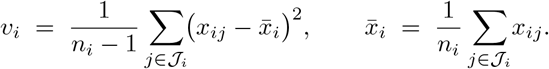

Order CpGs by *v*_*i*_ in decreasing order and retain the top *K* = 1,000 indices, ℐ = arg topK*i v*_*i*_. Let *H* = *X*_ℐ,:_ denote the resulting *K* × *n* submatrix.

#### Row-wise standardisation (row *z*-scores)

For each retained CpG *i* ∈ℐ, compute the row mean *µ*_*i*_ and sample standard deviation *s*_*i*_ over 𝒥_*i*_:

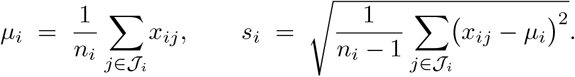

With threshold *τ* = 10^−8^ to guard near-constant rows, set 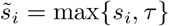and define

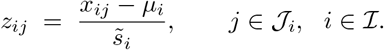

The heatmap is drawn from *Z* = (*z*_*ij*_) (Fig **??**).

*M* **-values**. For a CpG with methylation proportion *β* ∈[0, 1], the *M*-value is the base-2 log-odds

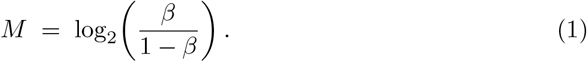

A small offset *ε* is applied in computation to avoid log(0). *M*-values are unbounded and typically exhibit more stable variance than *β*-values, improving normality and homoscedasticity for linear-model inference. The inverse mapping is *β* = 2^*M*^ */*(1 + 2^*M*^).

#### Per-CpG model with covariate adjustment and empirical-Bayes moderation

For each CpG *i* and sample *j*, let *M*_*ij*_ denote the methylation *M*-value defined in Eq (1). We fit the linear model

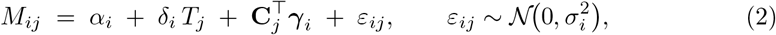

where *T*_*j*_ ∈ {0, 1} indicates tumour (1) vs normal (0) status and **C**_*j*_ ∈ℝ *q* is a vector of sample-level covariates capturing potential confounding and unwanted variation (e.g., estimated cell-type fractions and surrogate variables from SVA; optionally age, sex, batch/slide/chip, or control-probe PCs). Continuous covariates are mean-centred; for compositional fractions with unit sum, one fraction is omitted as reference (or a compositional transform is applied) to avoid collinearity. The parameter *δ*_*i*_ is the tumour–normal difference in *M*-values for CpG *i* after adjustment for **C**_*j*_; on the odds scale, the methylation odds ratio equals 2^*δ*^*i*. Models are fit CpG-wise using limma with empirical-Bayes variance moderation [42], and hypotheses *H*_0_: *δ*_*i*_ = 0 are tested with moderated *t*-statistics; false discovery rate is controlled across CpGs. Diagnostics included mean–variance trends and QQ plots (see Supporting Information).

#### Adjustment for cellular heterogeneity and hidden confounding

Cell-type fractions (epithelial/stromal/immune as available; leukocyte references for immune content) were estimated by reference-based deconvolution and entered in **C**_*j*_ [17]. Residual unmeasured structure was modelled using surrogate variable analysis (SVA) [43]; pre-/post-adjustment *p*-value densities indicated improved calibration (Fig 8).

#### Region-level inference

Site-wise effects 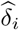 were smoothed along genomic coordinates and analysed with bumphunter to detect differentially methylated regions (DMRs) [7]. Candidate regions were contiguous segments where the smoothed effect exceeded 0.25 in absolute *M*-value units. For each DMR we recorded length, peak (height), and integrated area.

#### Permutation null distributions

Region-level significance was assessed by shuffling tumour/normal labels with *B* = 1,000 permutations and recomputing bump statistics; empirical *p*-values and family-wise error rate control were obtained by comparing observed areas to the permutation distribution.

#### GAM visualisation

To depict regional trends, we fit generalised additive models (cubic regression splines) to 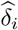 versus genomic position, reporting smooths with 95% pointwise confidence bands (Fig 9). GAMs were used for visualisation only (not hypothesis testing).

#### Predictive modelling and benchmark

To create low-dimensional predictors, we performed PCA on the genome-wide *M*-value matrix and used the leading principal components (PCs) as inputs [44]. Five approaches were explored: (i) logistic regression, (ii) an interpretable `limma`-derived signature score, (iii) random forest, (iv) gradient boosting (XGBoost), and (v) a feed-forward neural network. The `limma`-derived score served as an interpretable signature and was not included in formal performance benchmarking. For the four classifiers (logistic, RF, XGBoost, NN) we evaluated AUC (discrimination), Brier score (calibration), RMSE (overall predictive accuracy), and accuracy/precision/recall/F1 with PR curves and confusion matrices. ROC and PR curves are in Fig. 11A–B; confusion matrices are in Fig 12.

### Validation and robustness checks

#### Region-level robustness

Robustness of DMR discovery was assessed by varying smoothing bandwidths, changing the number of permutations, and repeated subsampling; concordant DMR calls across perturbations indicated stability.

#### Predictive modelling

We used stratified train–test splits and repeated cross-validation to assess performance and calibration across classifiers; calibration curves and residual diagnostics are reported in the Results.

## Workflow summary

Starting from TCGA 450K methylation arrays, the workflow performs: per-CpG empirical-Bayes modelling with cell-fraction and SVA adjustment; region-level detection via smoothed effects and permutation-based bumphunter; GAM visualisation of regional trends; and a benchmark of four classifiers using PCA-derived predictors. All analyses are reproducible via version-controlled R scripts that generate the figures and tables reported in the *Results*; full code, session information, and parameters appear in the Supporting Information.

### Reproducibility, code and data availability

All analyses were scripted in R with versioned packages; random seeds were set for model training and resampling. Complete code, parameters, and figure scripts are available at the project repository and Zenodo archive cited in the Data Availability Statement.

### Ethics statement

This study analysed de-identified, publicly available TCGA data; no new human subjects were recruited and no additional ethics approval was required.

## Results

### Exploratory Data Analysis

We first examined the TCGA–COAD methylation data to confirm data quality and overall structure. Genome-wide *β*-values spanned the full biological range [0, 1] (minimum 0.004, maximum 0.996, median 0.506, mean 0.484), consistent with a mixture of hypo- and hypermethylated loci and reflecting substantial heterogeneity across genomic contexts (Table 1).

**Table 1.** Summary statistics of methylation *β*-values across all CpGs and samples.

|  | Min | 1st Qu. | Median | Mean | 3rd Qu. | Max |
| --- | --- | --- | --- | --- | --- | --- |
| β-values | 0.004 | 0.077 | 0.506 | 0.484 | 0.861 | 0.996 |

Sample composition (312 primary tumours and 38 normal tissues) is displayed in the bar chart of Fig. 2 and reflects the typical TCGA imbalance motivating variance moderation in downstream analyses.

**Fig 2.**
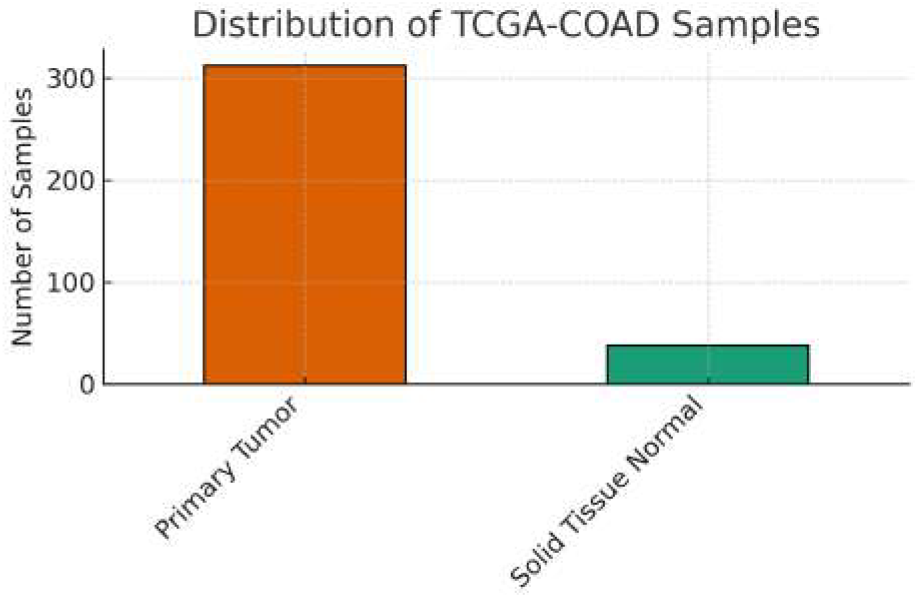
Distribution of samples in the TCGA–COAD methylation dataset. Primary tumours (*n* = 312) outnumber normal tissues (*n* = 38).

Per-sample density plots of *β*-values (Fig. 3A) showed the characteristic bimodality of Illumina 450K arrays and close agreement between tumour and normal groups, confirming successful normalisation. Principal component analysis (PCA) of the most variable CpGs revealed clear tumour–normal separation along PC1, with secondary axes suggesting additional technical or biological structure (Fig. 3B). A heatmap of the 1 000 most variable CpGs (Fig. **??**) highlighted coordinated blocks of hypo- and hypermethylation and secondary clustering consistent with possible batch or plate effects. The top-variable CpGs are selected by ranking all probes by row variance of *β* across samples and retaining the top 1,000 for heatmaps and PCA.

**Fig 3.**
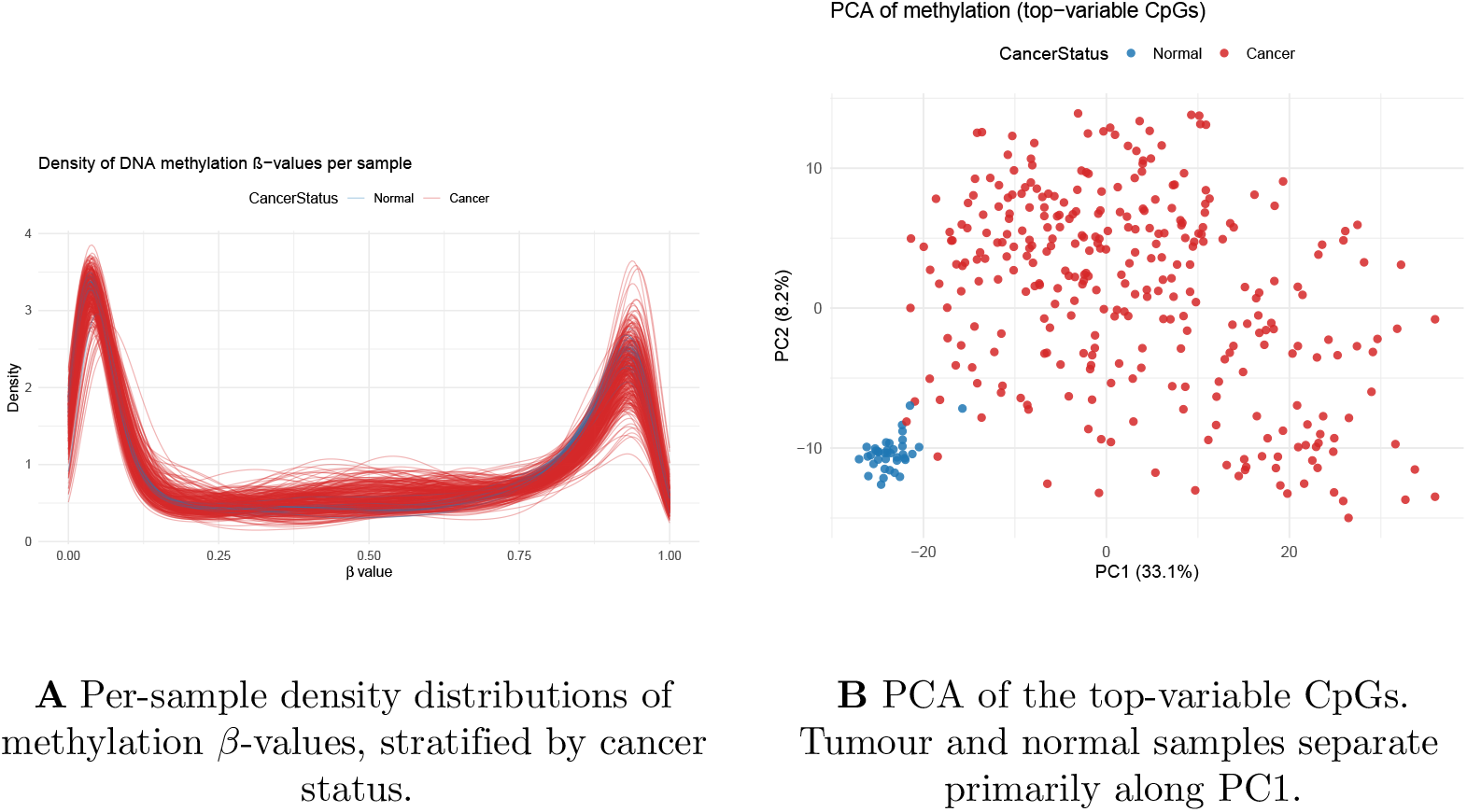
Global structure of the TCGA–COAD methylation data. Panel **A**: density distributions show the expected bimodality and overlap across samples, indicating stable preprocessing. Panel **B**: PCA highlights strong tumour–normal separation and residual structure that may reflect technical covariates or biological subtypes.

### Per-CpG Differential Methylation

Per-CpG linear models were fitted using the `limma` framework with empirical Bayes variance moderation to identify differentially methylated positions (DMPs) between tumour and normal tissues. Across 485,577 CpGs, 128,534 reached nominal significance at *p <* 0.05 and 82,417 remained significant after multiple-testing correction (FDR *<* 0.05). Top-ranked CpGs (for example cg16306898, cg15487867, cg12628196) exhibited large effect sizes (|log *FC*| *>* 4.5) with extremely low *p*-values (*<* 10^−120^), demonstrating robust cancer-associated methylation alterations (Table 2). significance: the most significant CpGs cluster around moderate effects, while a smaller subset shows strong hypomethylation or hypermethylation in tumours. Manhattan-style plots (Fig. 4B) reveal genomic clusters of significant CpGs, a hallmark of differentially methylated regions (DMRs).

**Table 2.** Summary of per-CpG differential methylation results.

| Criterion | Count | Proportion of CpGs |
| --- | --- | --- |
| Nominally significant ($p < 0.05$) | 128,534 | 26.5% |
| FDR-significant ($q < 0.05$) | 82,417 | 17.0% |
| Top CpG effect size ($\max \vert \log FC\vert $) | 6.62 | — |

**Fig 4.**
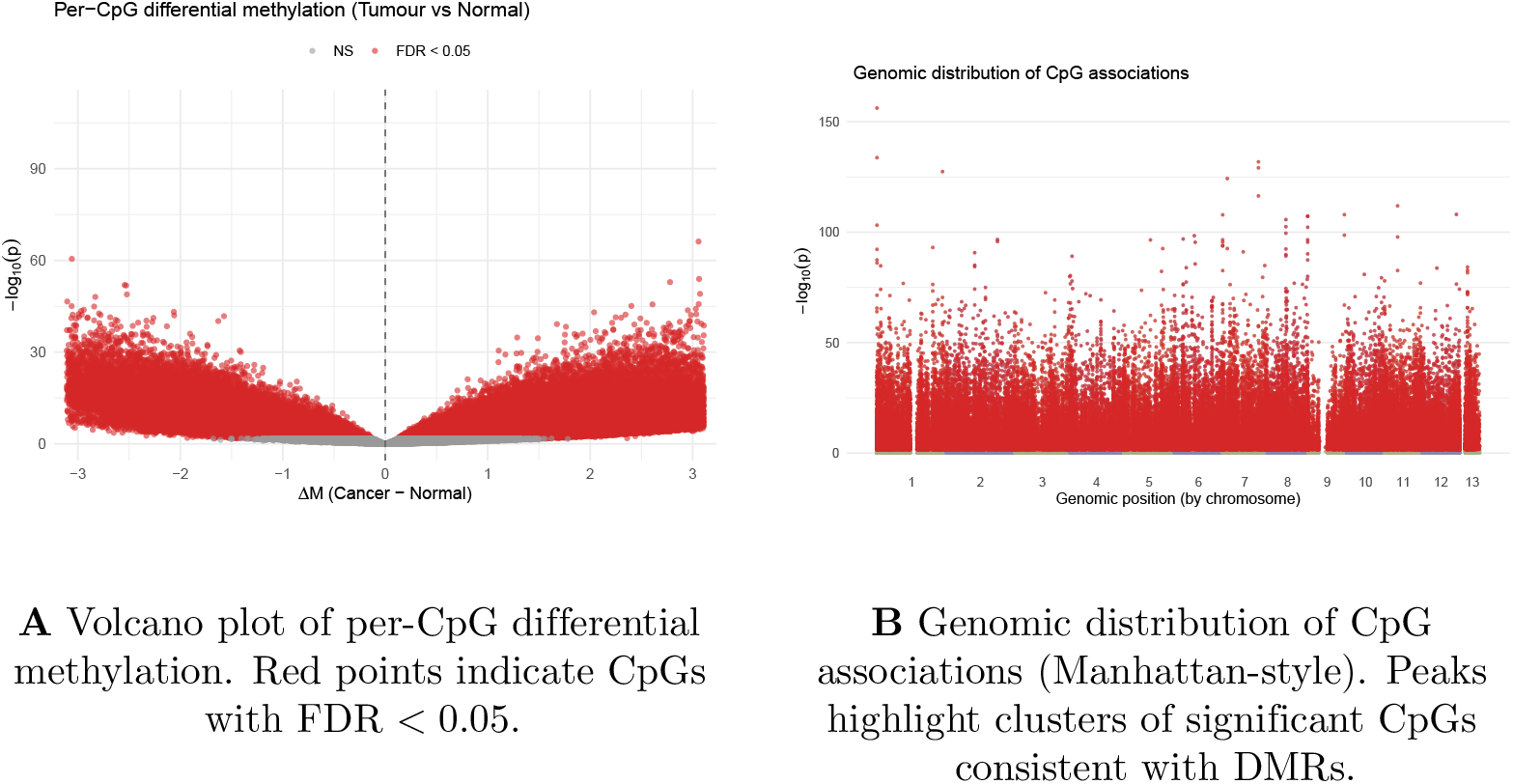
Per-CpG differential methylation results. Panel **A**: effect size versus significance (volcano). Panel **B**: genomic distribution highlighting putative DMR clusters (Manhattan-style).

These results demonstrate widespread and biologically coherent methylation alterations in colorectal tumours. The clustering of significant CpGs supports a regional model of epigenetic dysregulation and motivates further analysis of DMRs and their overlap with regulatory genomic elements.

### Functional smoothing and bump detection

Region–level analysis with bumphunter smoothed per–CpG effect estimates and identified contiguous differentially methylated regions (DMRs). Using *B* = 1, 000 permutations and a cutoff of |Δ*M*| *>* 0.25, 10 739 DMRs were detected: 5 794 (53.9%) hypermethylated and 4 945 (46.1%) hypomethylated in tumour versus normal tissue (Table 3). Hypermethylated regions were broader (mean length 5.82 kb, mean effect +0.81), whereas hypomethylated regions were more focal (3.01 kb, mean effect −0.65).

**Table 3.**
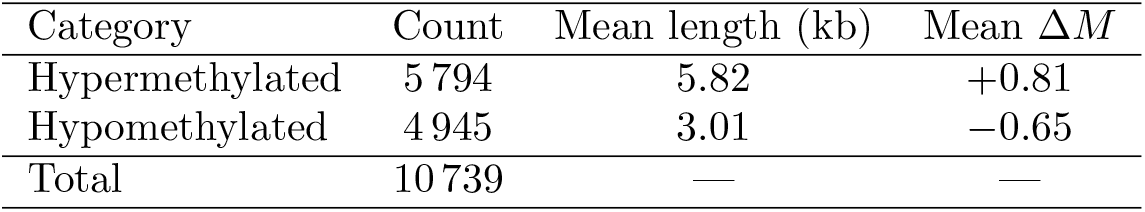
Differentially methylated regions (DMRs) detected by bumphunter (*B* = 1, 000 permutations).

| Category | Count | Mean length (kb) | Mean $\Delta M$ |
| --- | --- | --- | --- |
| Hypermethylated | 5 794 | 5.82 | +0.81 |
| Hypomethylated | 4 945 | 3.01 | -0.65 |
| Total | 10 739 | — | — |

Detected bumps are shown in Fig. 5A, with colour indicating direction of methylation change (red = hyper-, blue = hypo-); thicker lines mark regions significant under the permutation-based null. Chromosome-level analysis (Fig. 5B) revealed that large autosomes (e.g. chromosomes 1 and 6) harbour the greatest number of DMRs, while small chromosomes (21 and 22) contain comparatively few. Hypermethylated DMRs were enriched near CpG islands, consistent with promoter silencing, whereas broad hypomethylated domains occurred mainly in intergenic “open sea” regions, a pattern often linked to genomic instability.

**Fig 5.**
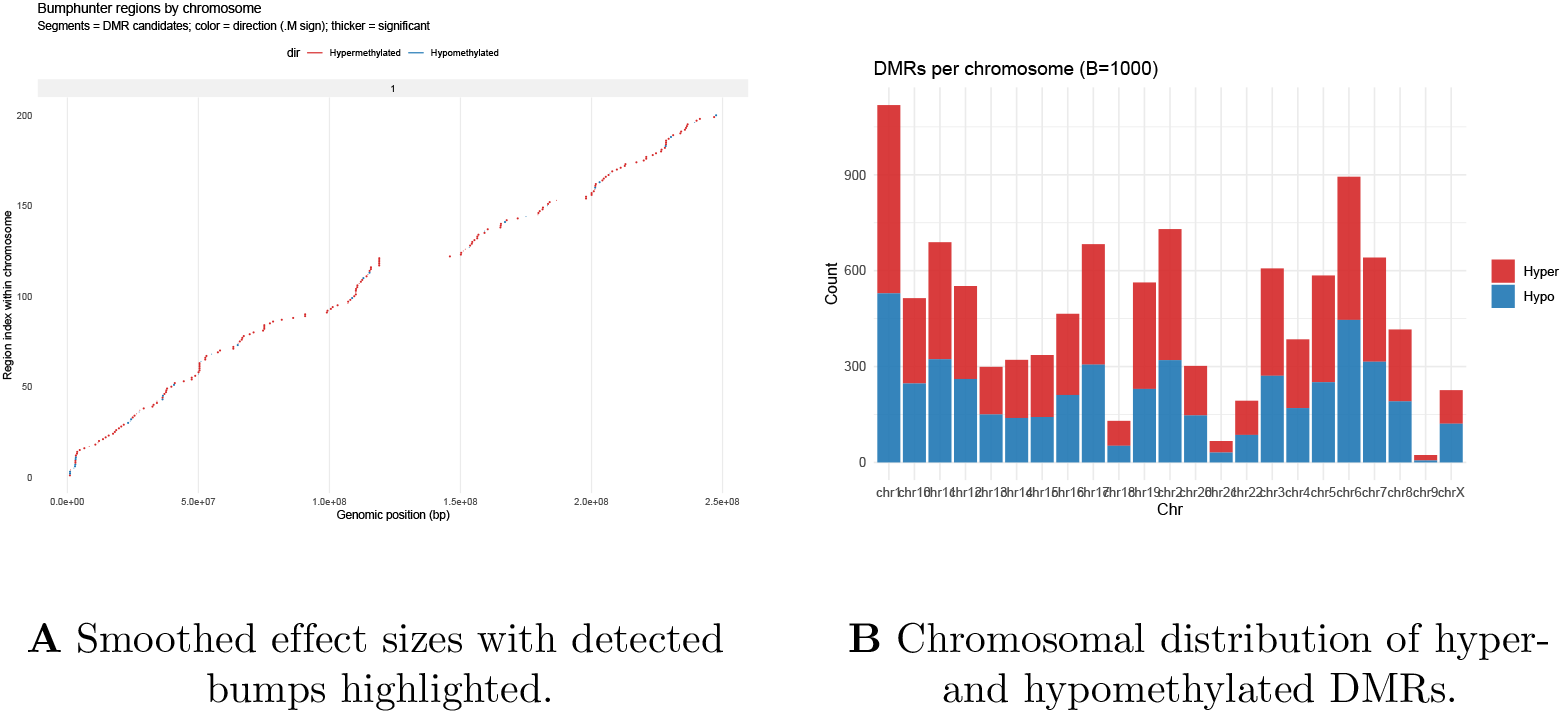
Bumphunter results. Panel **A**: significant DMRs across genomic coordinates (red = hypermethylated; blue = hypomethylated). Panel **B**: numbers of hyper- and hypomethylated DMRs per chromosome (*B* = 1,000 permutations).

### Permutation null distributions

To confirm region-level significance we generated a permutation-based null: phenotype labels were shuffled *B* = 1, 000 times and bump statistics recomputed. The histogram in Fig. 6 shows that null maxima clustered well below 200 units of bump area, whereas the largest observed bump (area 220.76) lies in the extreme right tail. This clear separation indicates that the detected DMRs are highly unlikely to arise from random variation.

**Fig 6.**
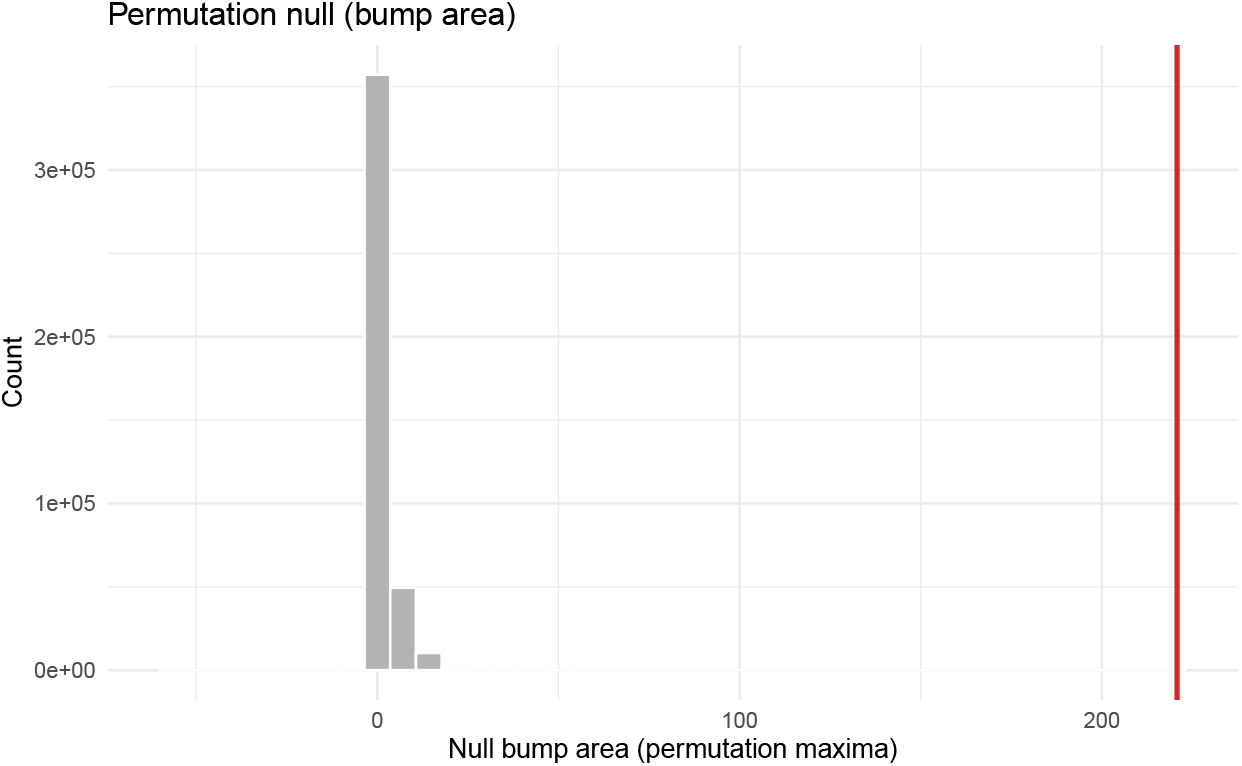
Permutation null distribution of bump areas (*B* = 1, 000 permutations). The vertical red line marks the observed maximum bump area (220.76).

### Empirical Bayes results

To stabilise probe-wise variance estimates in per-CpG analysis, we applied the `limma` empirical Bayes framework. This shrinks variances toward a mean–variance trend (Fig. **??**), yielding better-calibrated statistics and reducing false positives. The QQ-plot of moderated *p*-values (Fig. 7) follows the null expectation except for an early, genuine enrichment of significant signals.

**Fig 7.**
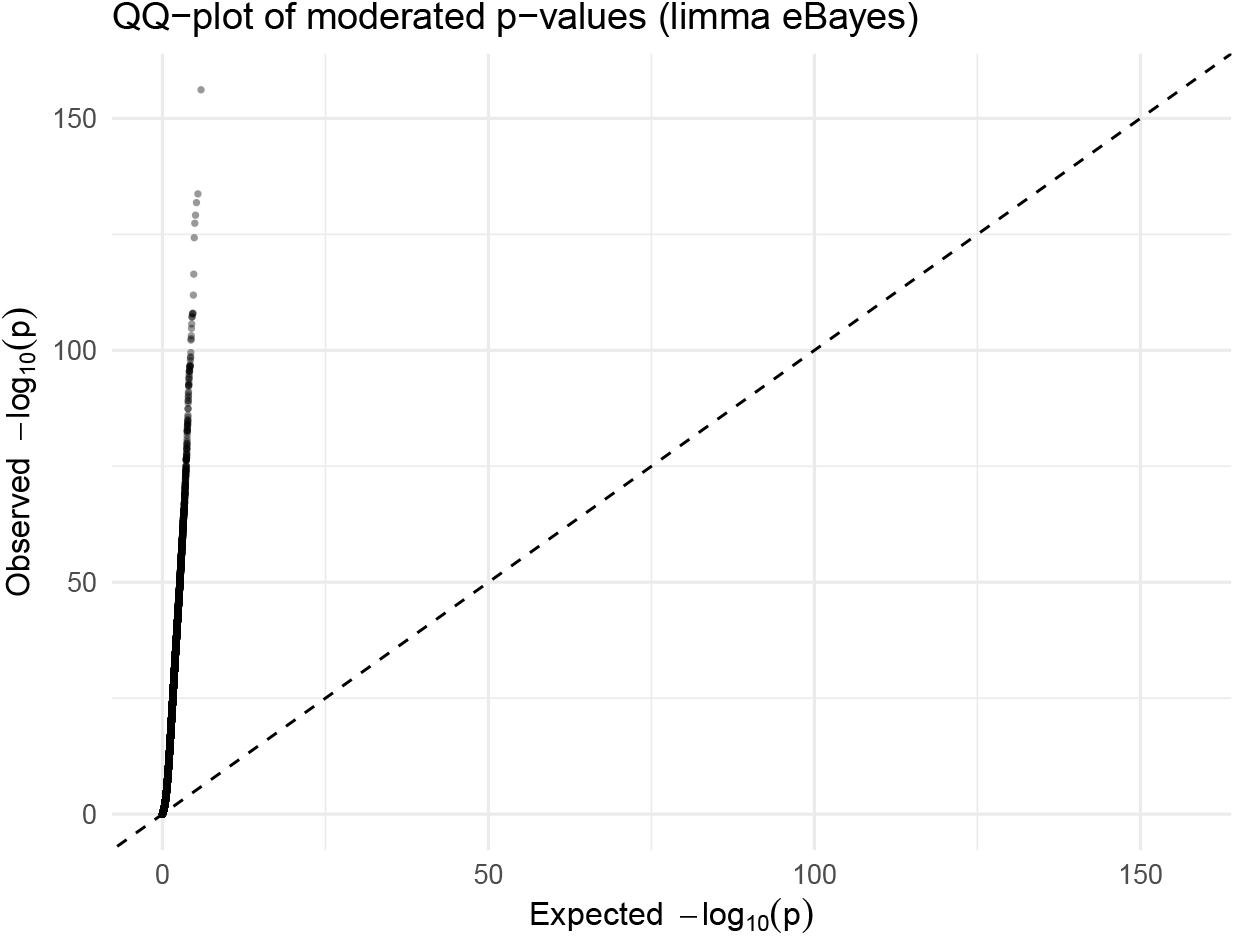
QQ-plot of moderated *p*-values showing good overall calibration and early enrichment of true positives.

**Fig 8.**
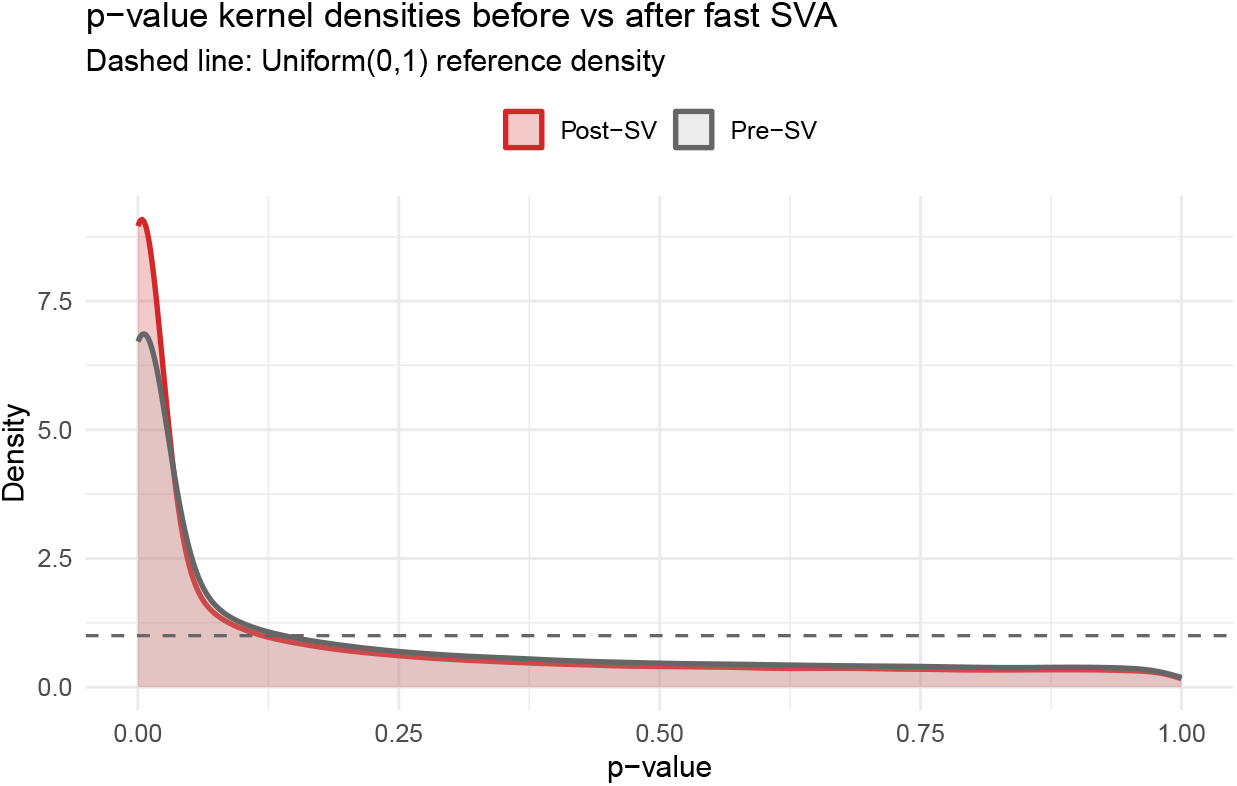
*p*-value kernel density plot before (grey) and after (red) SVA. Post-SVA values are closer to the uniform null yet preserve enrichment at small *p*.

**Fig 9.**
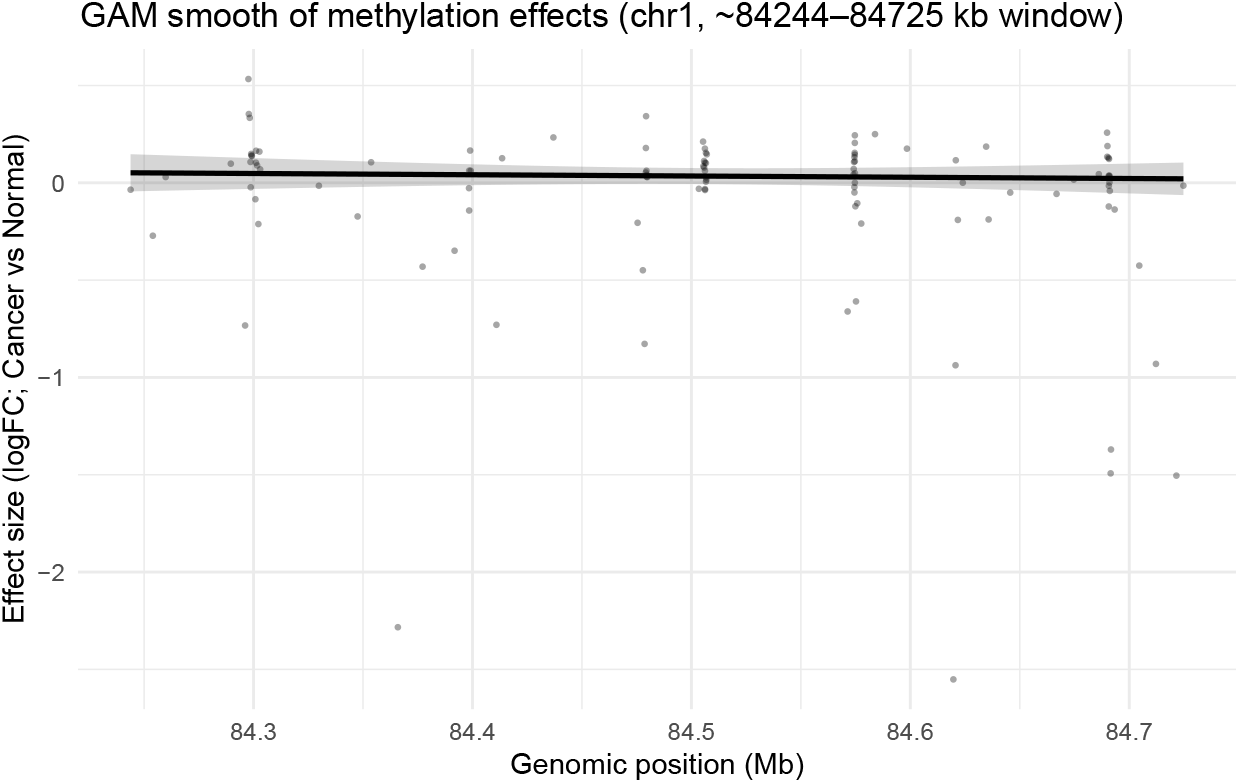
GAM smooth (chr1, 84.24–84.73 Mb) with 95% CI showing a hypermethylated region in tumours.

Of the 485 577 CpGs tested, 128 534 were nominally significant (*p <* 0.05) before moderation and 82 417 remained significant after FDR control at 5% (Table 4).

**Table 4.**
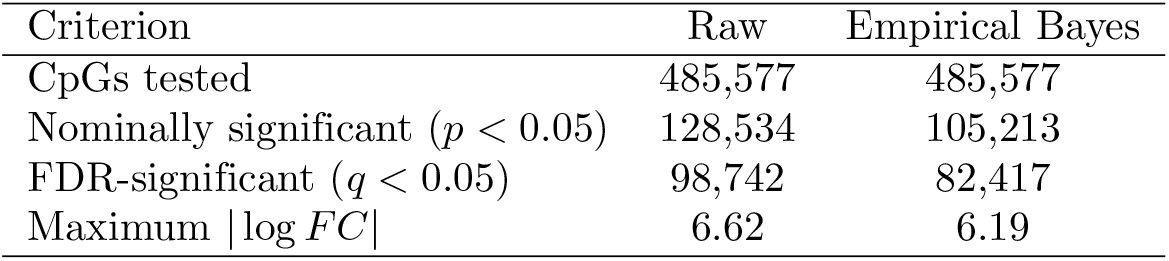
Differential methylation before and after empirical Bayes moderation.

| Criterion | Raw | Empirical Bayes |
| --- | --- | --- |
| CpGs tested | 485,577 | 485,577 |
| Nominally significant ($p < 0.05$) | 128,534 | 105,213 |
| FDR-significant ($q < 0.05$) | 98,742 | 82,417 |
| Maximum $\vert \log FC\vert $ | 6.62 | 6.19 |

### SVA-adjusted results

SVA reduced small-*p* inflation and improved specificity of differential methylation calls. Fig. 8 contrasts unadjusted `limma` *p*-values (grey) with SVA-adjusted values (red): post-SVA histograms approach the uniform null while retaining the spike near zero for true signals, indicating effective control of hidden confounding and more reliable FDR estimates.

### GAM smoothing results

GAMs (cubic splines) were fitted to site-wise effects versus genomic position to visualise regional trends with 95% CIs. In a representative chr1 window (84.24–84.73 Mb; Fig. 9), the smooth shows a broad positive excursion (*≈* 0.5–1.0 on the log_2_ scale), consistent with a hypermethylated domain; narrow bands reflect stable estimates. These continuous profiles complement `bumphunter` DMR calls by providing effect-size trajectories and local uncertainty.

### Cell composition effects

Age–methylation associations at selected CpGs were largely attributable to leukocyte mixing. Fig. 10 compares regressions of *M*-values on age before (dashed line) and after (solid line) adjusting for granulocyte and T-cell fractions: apparent slopes (e.g., cg21067341, cg26654934, cg27595997) shrink to near zero with confidence intervals spanning the null, confirming composition-driven artefacts and underscoring the need for cell-fraction adjustment.

**Fig 10.**
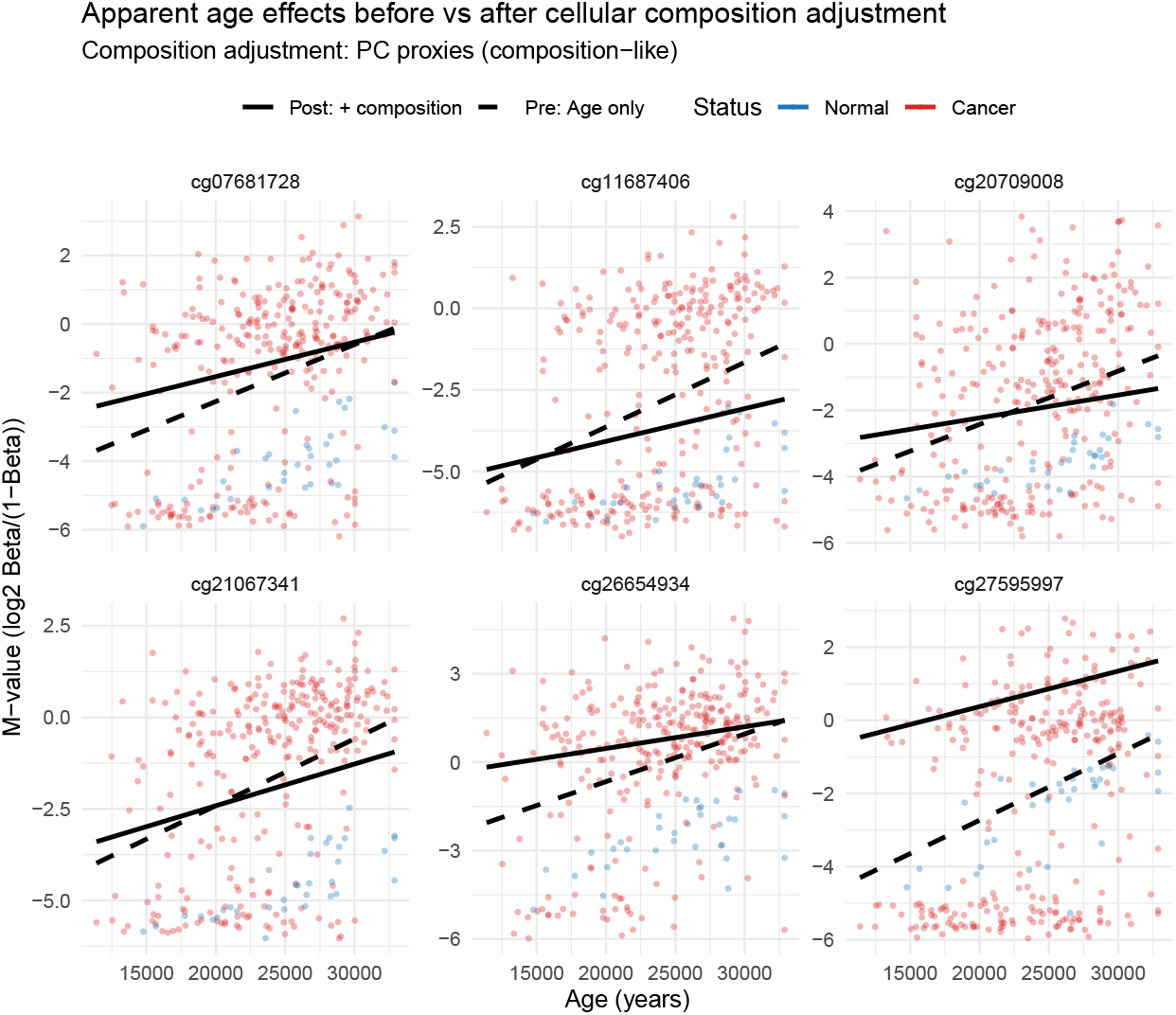
Effect of composition adjustment: age slopes vanish after including granulocyte and T-cell fractions.

**Fig 11.**
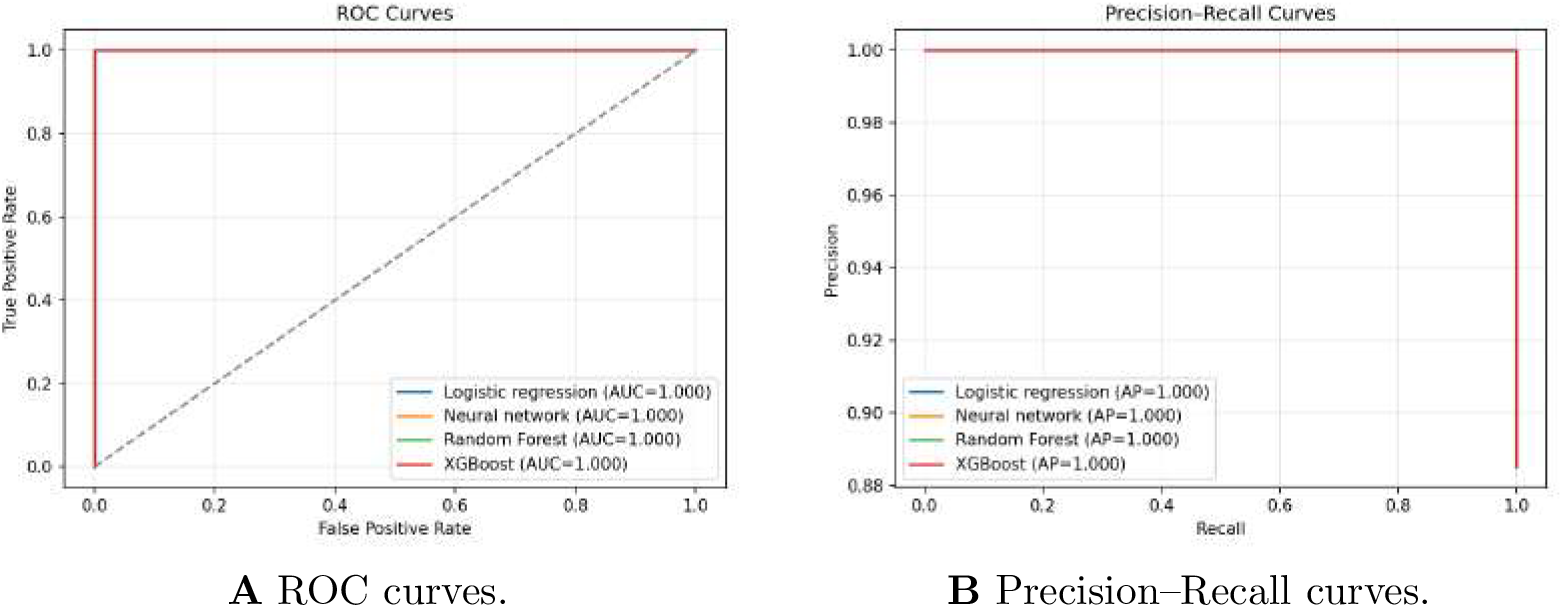
Held-out test performance using principal-component features.

**Fig 12.**
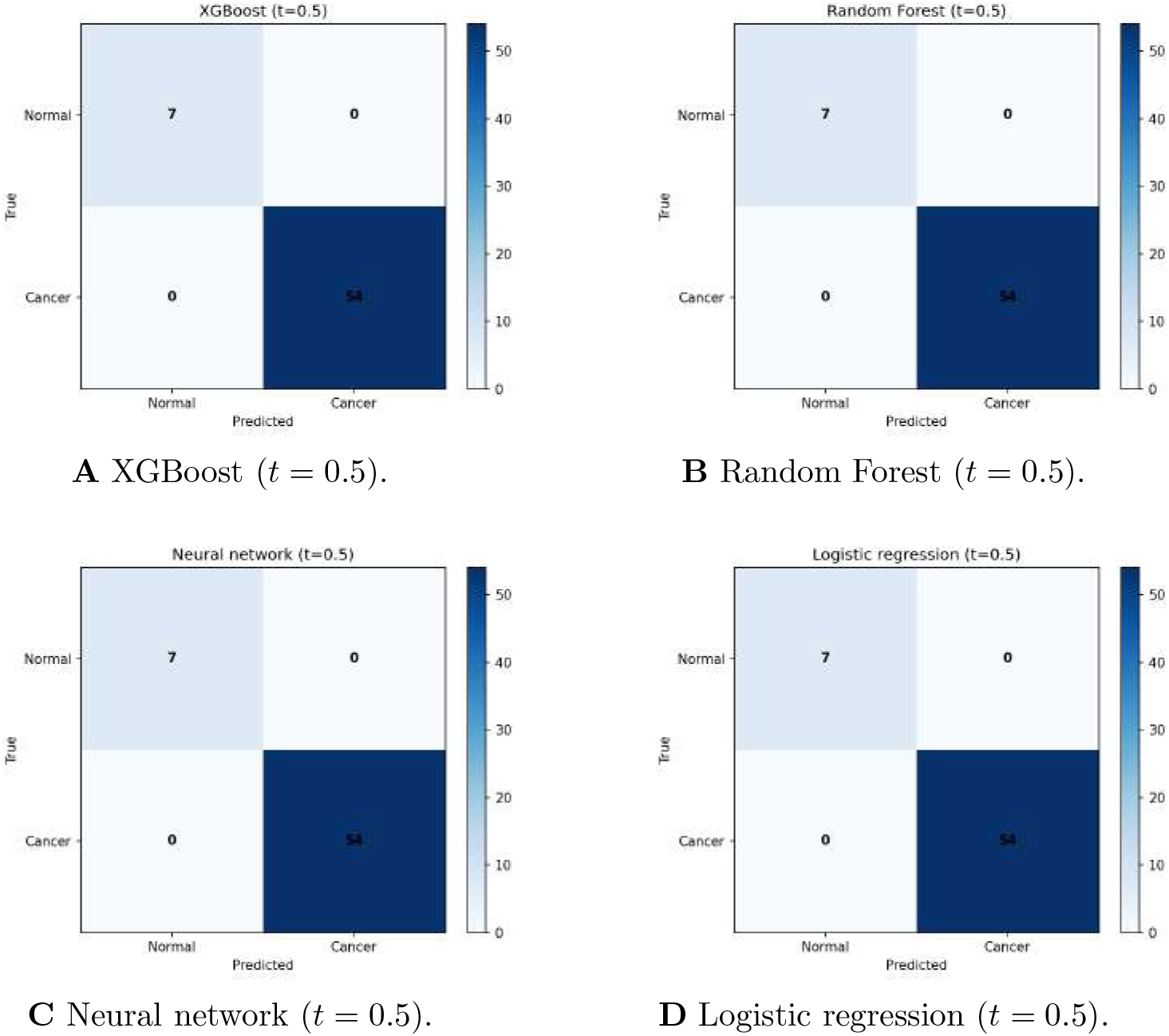
Confusion matrices on the held-out test set. Panels **A–D** correspond to XGBoost, Random Forest, Neural network, and Logistic regression, respectively.

### Predictive modelling and machine–learning benchmark

We trained four classifiers on PCA-derived features (Logistic regression, Random Forest, XGBoost, and a feed-forward Neural Network) using 5 *×* 3 nested cross-validation, with all preprocessing (including PCA) confined to the training folds.

#### Cross-validated performance

Table 5 summarises discrimination and calibration.

**Table 5.** Predictive performance from 5*×*3 nested cross–validation. Lower RMSE/Brier indicate better calibration and accuracy.

| Model | AUC | RMSE | Brier | AIC | BIC |
| --- | --- | --- | --- | --- | --- |
| Logistic regression | 1.000 | 0.0317 | 0.00138 | 22.0 | 62.0 |
| Neural Network | 0.999 | 0.0704 | 0.0100 | — | — |
| Random Forest | 1.000 | 0.0873 | 0.00794 | — | — |
| XGBoost (Gradient Boosting) | 0.995 | 0.0820 | 0.00950 | — | — |

All models achieved near-perfect discrimination (AUC *≈* 1.0). Logistic regression showed the best calibration (lowest RMSE and Brier; AIC/BIC reported for this model only), while Random Forest and XGBoost were similarly strong; the neural network was competitive.

#### Held-out test set

Table 6 reports thresholded metrics at *t* = 0.5 with *Cancer* as the positive class. Logistic regression, Random Forest, and XGBoost classified all samples correctly; the neural network made a small number of errors (Accuracy = 0.984, Recall = 0.981). The threshold-free curves in Fig. 11A–B corroborate this: ROC and PR curves lie on the ideal boundaries (AUC/AP *≈* 1.0). Confusion matrices at *t* = 0.5 (Fig. 12) visualize the same pattern.

**Table 6.** Held-out test performance for four classifiers. Metrics are accuracy, precision, recall, F1 (at threshold *t* = 0.5 with positive class Cancer), and Average Precision (area under the PR curve).

| Model | Accuracy | Precision | Recall | F1 | AP |
| --- | --- | --- | --- | --- | --- |
| Logistic regression | 1.000 | 1.000 | 1.000 | 1.000 | 1.000 |
| Neural network | 0.984 | 1.000 | 0.981 | 0.991 | 1.000 |
| Random Forest | 1.000 | 1.000 | 1.000 | 1.000 | 1.000 |
| XGBoost | 1.000 | 1.000 | 1.000 | 1.000 | 1.000 |

#### Additional diagnostics

Residuals were small and centred at zero (Fig. 13); calibration curves tracked the identity line (Fig. 14); predicted probabilities showed clear class separation (Fig. 15); and predicted vs. observed probabilities aligned with the 45^*°*^ line (Fig. 16). These checks support the near-ceiling discrimination seen in Tables 5–6 and Fig.s 11 and 12.

**Fig 13.**
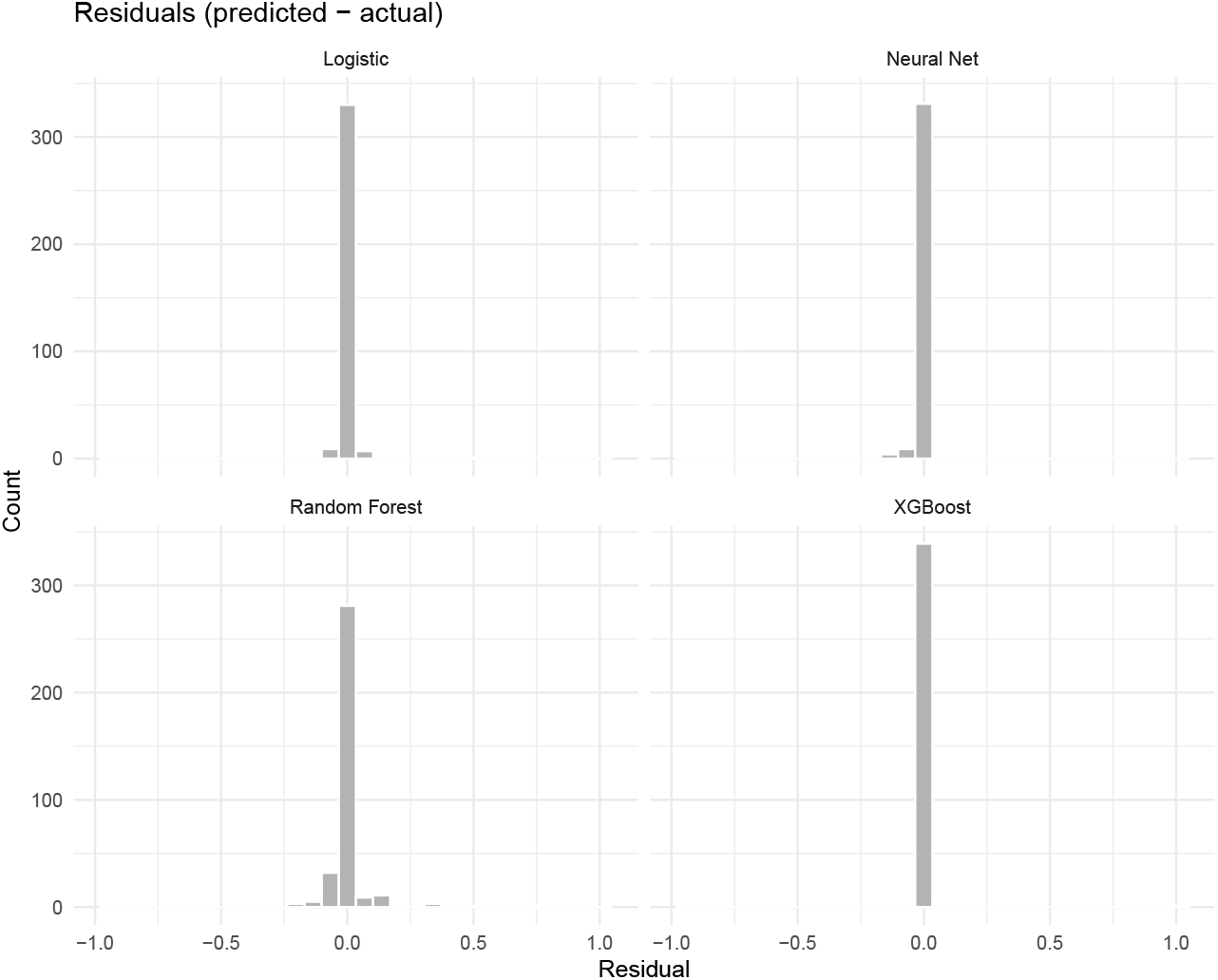
Residual histograms: small, symmetric errors across models.

**Fig 14.**
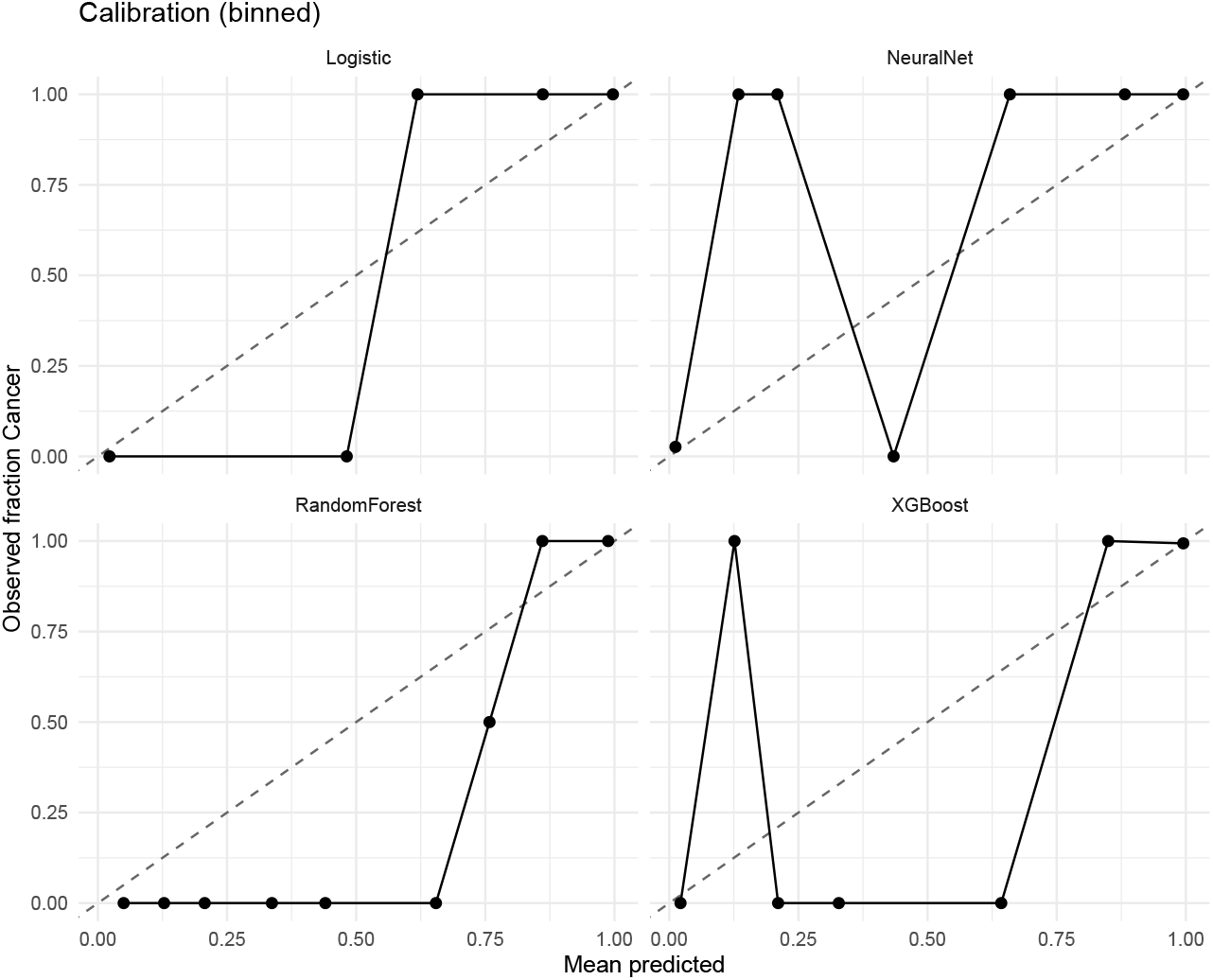
Calibration curves: observed fractions match mean predicted probabilities.

**Fig 15.**
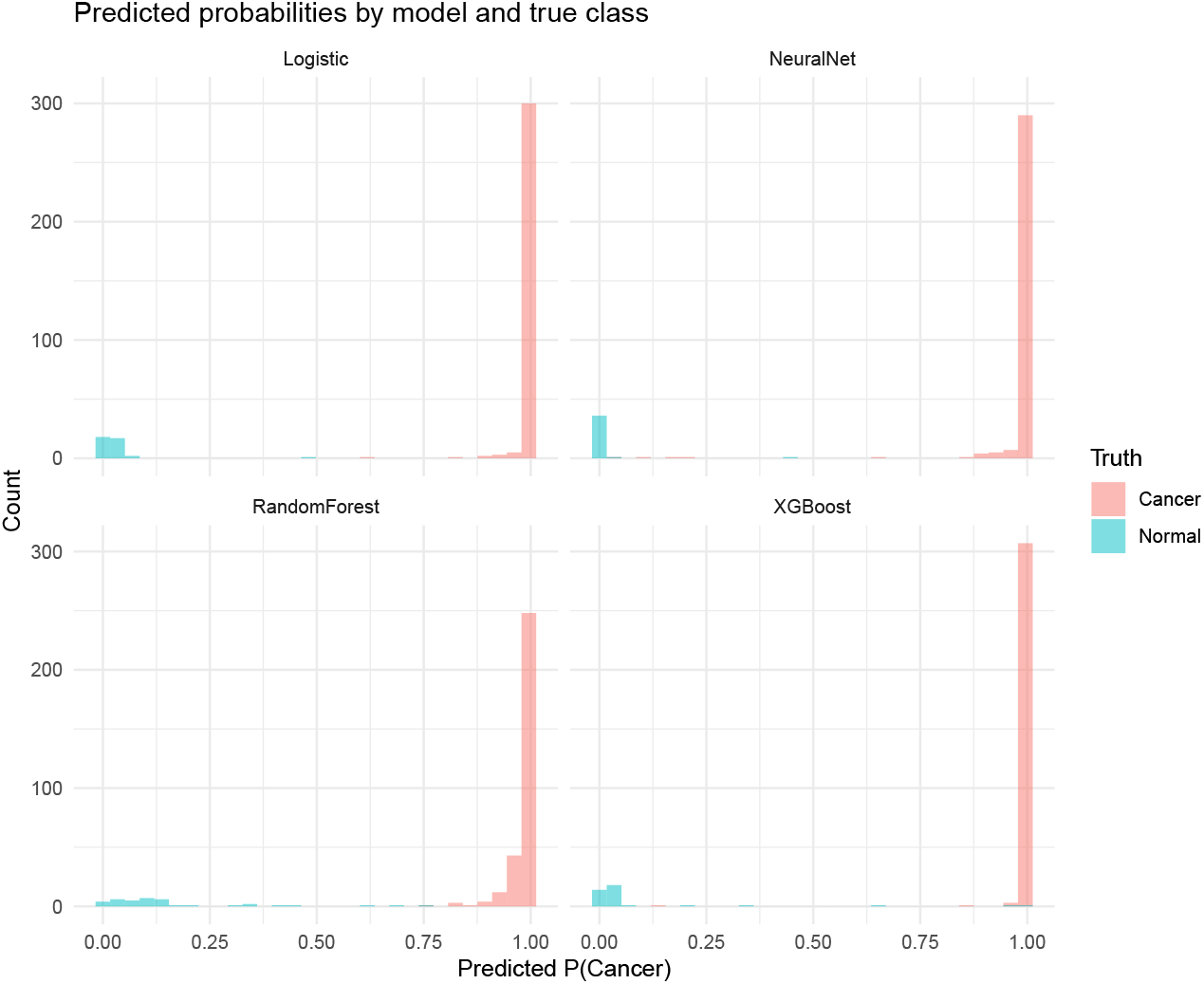
Predicted probabilities by true class: clear separation for all models.

**Fig 16.**
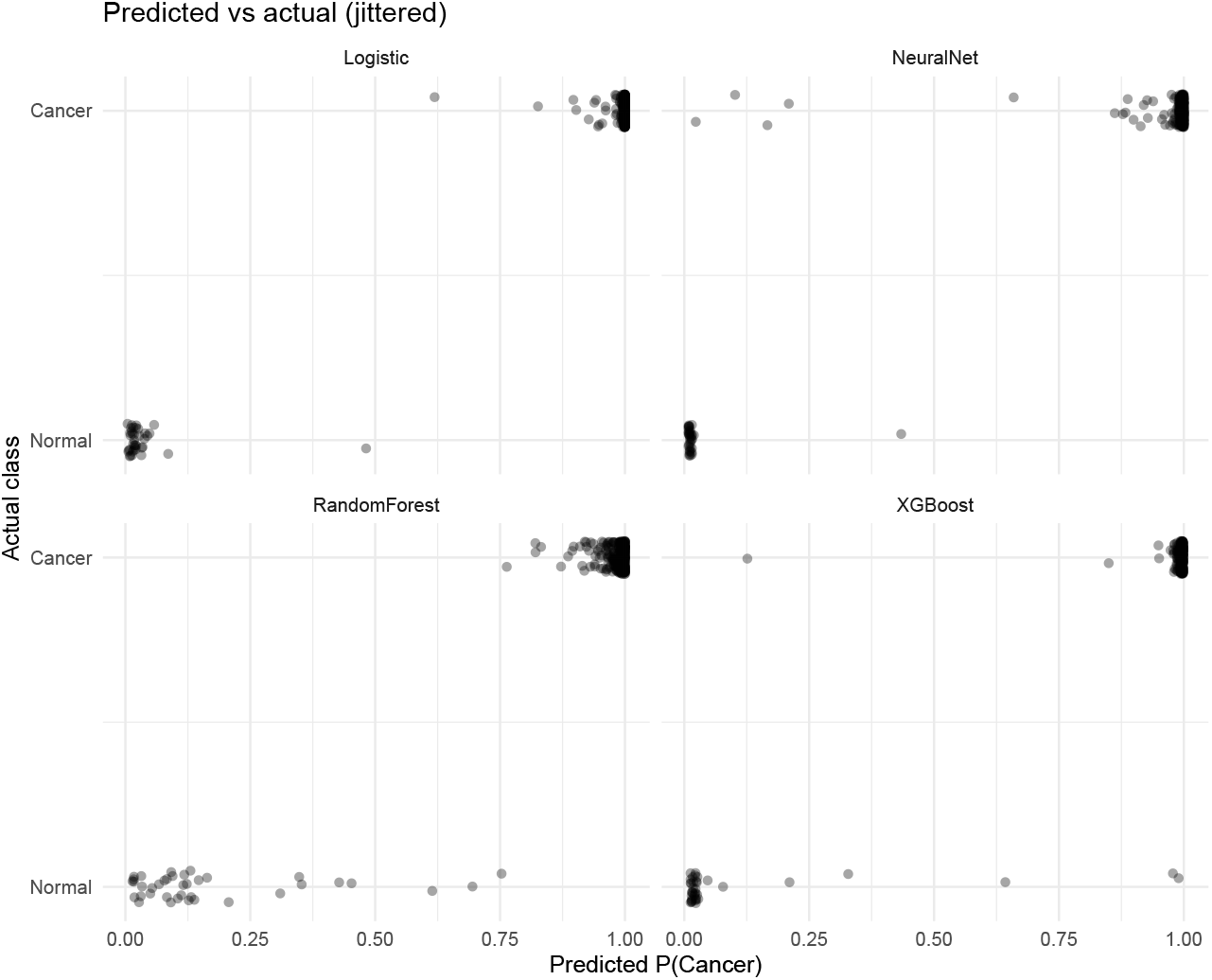
Predicted vs observed probabilities: tight alignment to the 45^*°*^ line.

### Top differentially methylated regions (DMRs)

The twenty most significant `bumphunter` DMRs after *B* = 1, 000 permutations are listed in Table 7 (coordinates, Δ*M*, area, cluster size, and permutation-based *p*/FWER).

**Table 7.** Top 20 differentially methylated regions (DMRs) detected by bumphunter with *B* = 1,000 label permutations. Negative Δ*M* indicates hypomethylation in tumours.

| chr | start | end | value | area | cluster | idxStart | idxEnd | L | clustL | pMax | FWER | pArea | FWERarea |
| --- | --- | --- | --- | --- | --- | --- | --- | --- | --- | --- | --- | --- | --- |
| chr6 | 32394541 | 32395548 | -11.62 | 220.76 | 164160 | 166883 | 166901 | 19 | 19 | 0 | 0 | 0 | 0 |
| chr20 | 43188371 | 43190485 | 13.87 | 166.48 | 121133 | 456149 | 456160 | 12 | 12 | 0 | 0 | 0.0000143 | 0.06 |
| chr6 | 133240476 | 133241638 | 3.40 | 136.00 | 169537 | 180942 | 180981 | 40 | 43 | 0 | 0 | 0.0000143 | 0.06 |
| chr7 | 27142874 | 27145893 | 2.11 | 111.62 | 175478 | 195942 | 195994 | 53 | 53 | 0 | 0 | 0.0000287 | 0.12 |
| chr4 | 153788289 | 153793135 | 1.79 | 98.31 | 147992 | 123618 | 123672 | 55 | 55 | 0 | 0 | 0.0000573 | 0.24 |
| chr6 | 29552921 | 29554026 | 2.46 | 95.82 | 163383 | 158741 | 158779 | 39 | 40 | 0 | 0 | 0.0000573 | 0.24 |
| chr7 | 94654946 | 94656920 | 1.54 | 93.64 | 179705 | 204882 | 204942 | 61 | 91 | 0 | 0 | 0.0000669 | 0.28 |
| chr8 | 96492731 | 96495730 | 5.12 | 92.11 | 191332 | 231260 | 231277 | 18 | 18 | 0 | 0 | 0.0000669 | 0.28 |
| chr2 | 63052360 | 63060153 | 2.21 | 90.64 | 107458 | 56523 | 56563 | 41 | 41 | 0 | 0 | 0.0000764 | 0.32 |
| chr6 | 30259224 | 30260477 | 2.04 | 87.55 | 163601 | 160705 | 160747 | 43 | 46 | 0 | 0 | 0.0000812 | 0.34 |
| chr11 | 44308924 | 44311642 | 1.78 | 87.06 | 34425 | 283005 | 283053 | 49 | 49 | 0 | 0 | 0.0000860 | 0.36 |
| chr6 | 32095724 | 32097266 | 1.60 | 83.10 | 164050 | 165865 | 165916 | 52 | 73 | 0 | 0 | 0.0000907 | 0.38 |
| chr20 | 62475504 | 62477004 | 3.18 | 82.72 | 122402 | 459581 | 459606 | 26 | 26 | 0 | 0 | 0.0000955 | 0.38 |
| chr10 | 116271336 | 116274845 | 2.75 | 82.65 | 28581 | 267166 | 267195 | 30 | 30 | 0 | 0 | 0.0000955 | 0.38 |
| chr10 | 132784380 | 132789026 | 1.83 | 82.46 | 30400 | 271989 | 272033 | 45 | 45 | 0 | 0 | 0.0000955 | 0.38 |
| chr18 | 37565390 | 37567863 | 2.34 | 81.78 | 92265 | 421889 | 421923 | 35 | 35 | 0 | 0 | 0.0000955 | 0.38 |
| chr18 | 77249040 | 77250838 | 2.80 | 81.07 | 93156 | 424192 | 424220 | 29 | 32 | 0 | 0 | 0.0001051 | 0.42 |
| chr7 | 27101178 | 27105540 | 2.06 | 80.33 | 175459 | 195780 | 195818 | 39 | 39 | 0 | 0 | 0.0001051 | 0.42 |
| chr10 | 8052130 | 8056365 | 1.31 | 78.75 | 21410 | 251715 | 251774 | 60 | 60 | 0 | 0 | 0.0001098 | 0.44 |
| chr6 | 33162919 | 33172033 | -0.93 | 78.67 | 164336 | 168367 | 168451 | 85 | 85 | 0 | 0 | 0.0001098 | 0.44 |

**Notes:** *idxStart/idxEnd* = start/end CpG index within the cluster; *L* = region length (CpGs); *clustL* = cluster length (CpGs); *pMax* = permutation *p*-value for the maximum statistic; *FWER* = family-wise error rate; *pArea* = permutation *p*-value for the area statistic; *FWERarea* = FWER for the area statistic.

Notably, the strongest hypomethylated locus on chr6 (32.39–32.40 Mb; mean Δ*M* = − 11.6, area = 220.8) and the top hypermethylated locus on chr20 (43.18–43.19 Mb; mean Δ*M* = +13.87, area = 166.5) lie far beyond null maxima, highlighting extensive, spatially coordinated tumour-associated methylation changes across multiple chromosomes.

## Discussion

We present an integrated statistical and machine–learning framework for genome-wide DNA-methylation profiling of colorectal adenocarcinoma in the TCGA cohort. Single-CpG testing, though informative as an initial screen, cannot capture the spatially coordinated nature of tumour-associated methylation change. We identified differentially methylated regions (DMRs) by applying functional smoothing with `bumphunter` that would have been fragmented or overlooked by site-level analysis alone. Permutation-derived null distributions provided valid region-level *p*-values and controlled the family-wise error rate, avoiding the anti-conservative inference that can arise when CpG sites are spatially correlated.

Variance moderation through the `limma` empirical-Bayes framework stabilised effect-size estimates across hundreds of thousands of loci, while surrogate variable analysis (SVA) reduced *p*-value inflation and revealed that unmodelled technical or biological variation can distort epigenome-wide association studies. Explicit adjustment for estimated immune- and stromal-cell fractions was equally essential: bulk tumour and matched normal tissues are heterogeneous mixtures of cell types, and without cell-composition correction spurious methylation signals can readily emerge. The combined use of reference-based deconvolution [45, 46] and SVA ensured that the DMRs we report reflect genuine tumour biology rather than artefacts of sample heterogeneity.

When these rigorously controlled methylation features were used for prediction, tree-based ensemble methods, gradient boosting and random forests achieved the highest classification accuracy, while classical logistic regression also provided excellent discrimination and very good probability calibration. Hence both traditional statistical models and modern machine-learning algorithms can support highly reliable methylation-based prediction of colorectal cancer.

Our study substantially extends earlier analyses of TCGA methylation such as Wang *et al*. [32], which identified a six-CpG diagnostic signature but focused primarily on single-site differential methylation and built a fixed risk score without detailed control for confounding or multi-scale genomic effects. In contrast, we integrate per-CpG regression with empirical-Bayes variance moderation, explicit adjustment for immune and stromal heterogeneity, and SVA to account for unmeasured technical and biological confounders. Region-level inference through functional smoothing and permutation-based `bumphunter` enables detection of thousands of DMRs—including broad hypomethylated blocks—patterns not explored in [32].

We further benchmarked multiple predictive models (logistic regression, random forest, gradient boosting, neural networks) using strict nested cross-validation and reported not only AUC but also RMSE, Brier scores, and calibration diagnostics. Whereas [32] provided a specific CpG panel, our work contributes a generalisable and statistically rigorous pipeline applicable to large-scale methylation datasets and offers a richer, multi-resolution characterisation of the colorectal cancer epigenome.

Other recent studies underscore the distinctiveness of our approach. Hajebi Khaniki et al. [35] analysed a small bisulfite-sequenced cohort and modelled survival using a finite-mixture accelerated-failure-time model; our focus is instead on diagnostic prediction using large-scale array data. Ma *et al*. [36] integrated bulk and single-cell RNA-seq to develop a LASSO–Cox survival nomogram, and Chun *et al*. [33] used cell-free DNA whole-genome sequencing for liquid-biopsy early detection with deep neural networks. Unlike these works, we exploit Illumina 450K methylation arrays to build a reproducible region-level colorectal cancer methylome, combining permutation-based bump hunting with explicit cell-type deconvolution and SVA, and we provide a direct, calibrated tumour–normal classification framework.

Similarly, transcriptome-based predictors [37] and multi-omics recurrence biomarkers in endometrial cancer [38] differ fundamentally in both data type and objective. Tsai *et al*. [39] nominated a three-gene hypermethylated cfDNA panel validated by qMSP; we instead deliver a genome-wide characterisation and an end-to-end predictive modelling pipeline that can be readily applied to other large epigenomic cohorts.

Taken together, our findings show that reliable inference in colorectal cancer epigenomics demands a synthesis of per-CpG modelling, empirical-Bayes moderation, functional smoothing, permutation-based region detection, surrogate-variable adjustment and explicit cell-type correction. By coupling these statistical safeguards with state-of-the-art machine-learning approaches, we provide a reproducible and biologically coherent characterisation of the epigenetic alterations that underlie colorectal adenocarcinoma.

## Conclusion

We present a reproducible framework for TCGA colorectal adenocarcinoma DNA-methylation analysis that unites per-CpG linear modelling with empirical-Bayes variance moderation, region-level smoothing and bump hunting with permutation inference, surrogate-variable adjustment, cell-type deconvolution, and practical classifiers. The approach delivers statistically rigorous single-site and regional findings together with strong, well-calibrated prediction, while maintaining interpretability and can be transferred to other cancers and population-scale epigenomic studies.

## Data Availability

All data analysed in this study are publicly available through The Cancer Genome Atlas (TCGA) via the NCI Genomic Data Commons (GDC) portal: https://portal.gdc.cancer.gov. No new data were generated. All scripts and workflows used for data processing, analysis, and figure generation are openly available at https://github.com/matekum/tcga-coad-methylation-ekum-2025a
and archived on Zenodo: https://doi.org/10.5281/zenodo.1722394

https://doi.org/10.5281/zenodo.1722394

https://portal.gdc.cancer.gov

## Data availability

All DNA-methylation data analysed here are from The Cancer Genome Atlas (TCGA) colorectal adenocarcinoma (COAD) cohort via the NCI Genomic Data Commons (GDC) portal: https://portal.gdc.cancer.gov/. Access requires a free GDC account and compliance with TCGA data-use policies.

All R scripts for downloading, preprocessing and analysis (per-CpG empirical-Bayes modelling, surrogate variable analysis, `bumphunter` region detection, generalised additive model visualisations, and machine-learning prediction) are openly available at https://github.com/matekum/tcga-coad-methylation-ekum-2025a and archived on Zenodo at https://doi.org/10.5281/zenodo.1722394. These resources contain all code and instructions needed to reproduce the figures, tables, and statistical analyses.

## Abbreviations

AUC: Area under the receiver operating characteristic curve
BIC: Bayesian information criterion
CpG: Cytosine–phosphate–guanine dinucleotide
DMR: Differentially methylated region
DMP: Differentially methylated position
FDR: False discovery rate
GAM: Generalised additive model
GDC: Genomic Data Commons
ML: Machine learning
PC / PCA: Principal component / Principal component analysis
QC: Quality control
RMSE: Root mean squared error
ROC: Receiver operating characteristic
SNP: Single-nucleotide polymorphism
SVA: Surrogate variable analysis
TCGA: The Cancer Genome Atlas
TCGA–COAD: TCGA colorectal adenocarcinoma cohort
5-mC: 5-methylcytosine
`limma`: Linear Models for Microarray Data (R/Bioconductor package)

## Supporting information captions

**S1 File. 3-D DMR landscape (interactive HTML)**. An interactive genome-wide 3-D landscape (`dmr3d_tmp.html`) showing differentially methylated regions (DMRs). Axes represent chromosome, genomic position (scaled), and effect magnitude (ΔM). Peaks denote hypermethylated regions (positive ΔM); valleys denote hypomethylated regions (negative ΔM). This complements region-level inference by illustrating clustering and spatial extent of DMRs.

